# Differences in rates of uptake of NICE clinical guidelines between Type 1 diabetes mellitus (T1DM) and Type 2 diabetes mellitus (T2DM) as evidenced by National Diabetes Audit of England and Wales

**DOI:** 10.1101/2020.08.05.20168914

**Authors:** Robert C Hayward, John Watkins, Cono Ariti

## Abstract

**OBJECTIVES:** To investigate whether there were important differences in uptake of NICE clinical guidelines according to diabetes mellitus type in England and Wales from 2013 to 2018, test the hypothesis that guidelines are more successfully implemented with T2DM than T1DM, and explore possible explanations for differences.

**DESIGN:** Retrospective cross-sectional analyses of aggregated patient level data from the National Diabetes Audit (NDA) dataset owned by NHS Digital and commissioned by Healthcare Quality Improvement Partnership.

**SETTING:** Diabetes specialist services, primary care (GP surgeries submitting NDA data) across England and Wales.

**PARTICIPANTS:** 1 739 175 patients with diabetes aged ≥20 in England and Wales in 2013-14, 1 871 320 individual patients in 2014-15, 2 688 106 individual patients in 2015-16, 3 095 275 individual patients in 2016-17, and 3 357 055 individual patients in 2017-18.

**INTERVENTIONS:** Recommended care for diabetes mellitus as outlined in relevant NICE guidelines and delivered by either specialist clinicians or GPs.

**MAIN OUTCOME MEASURES:** The recorded attainment of NICE treatment targets: HbA1c levels =<7.5% (58.5 mmol/mol), blood pressure <140/80mmHg, blood cholesterol <5mmol/l; and clinical processes: at least annual monitoring of HbA1c, blood pressure, cholesterol, albumin:creatinine ratio, smoking status, Body Mass Index.

**RESULTS:** Annual collections (2013-14, 2014-15, 2015-16, 2016-17 and 2017-18) were individually analysed, testing associations between diabetes type and attainment of clinical targets or processes using a multivariable logistic regression model, adjusted for age and sex. Increased odds of meeting clinical targets if patients had T2DM compared with T1DM was consistent across the five years, except for cholesterol levels <5mmol/l where T2DM patients had lower odds (all associations p<.0001). Greatest differences in all five years between T1DM and T2DM was observed with patients meeting the HbA1c=<7.5% target, the largest being in 2015-16 (Odds Ratio 3.43, 95% confidence interval 3.39 to 3.47).

**CONCLUSIONS:** The differences between T1DM and T2DM in HbA1c target attainment is key and potentially reflects challenges of managing T1DM with insulin but suggests a point of focus for that patient population. Other important elements for consideration could be specific setting for delivery (primary care versus secondary care) and duration of illness.

WHAT IS ALREADY KNOWN ON THIS TOPIC
NHS Digital publishes reports on the NDA every year, however much of this is descriptive rather than analytical. Very few published studies have explored NDA data and none have particularly compared clinical outcomes of T1DM and T2DM.

WHAT THIS STUDY ADDS
Differences between age structures of T1DM and T2DM patient groups are adjusted and success in meeting NICE clinical processes and treatment targets are compared. This study highlights the different challenges faced by these different groups and conditions and raises questions about the suitability of applying identical targets to the diabetes subtypes given aetiological, clinical and therapeutic differences.

## INTRODUCTION

There are many types of diabetes mellitus (a clinical syndrome characterised by hyperglycaemia), predominantly diagnosed as Type 1 diabetes mellitus (T1DM) or Type 2 diabetes mellitus (T2DM), depending on the clinically understood aetiology. In 2020, it is estimated that UK prevalence of diabetes is around 4.8 million, with 4.1 million of those living in England^1,2^. PHE’s prevalence model estimates total (diagnosed and undiagnosed) diabetes prevalence for people aged ≥16 years in England as 8.5%. Of these, approximately 8% have T1DM with most of the remainder having T2DM^3^. Overall prevalence in England is higher in men (9.6%) than women (7.6%), and higher in people from South Asian and black ethnic groups (15.2%) compared with those from white, mixed or other ethnic groups (8.0%) (Public Health England, 2016). Overall, diabetes prevalence increases as age increases, and there is significant local variation in prevalence rates^1^. There are health inequalities considerations for diabetes, as there is evidence that the most deprived quintile are 1.5 times more likely to develop diabetes than the least deprived quintile^4^. Deprivation is only associated with T2DM: perhaps unsurprising as T1DM incidence is not thought to be affected by lifestyle factors^2^.

For the UK, the body responsible for the development of clinical or medical guidelines to improve and standardise clinical practice is the National Institute for Health and Care Excellence (NICE). Clinical guidelines are “systematically developed statements to assist practitioner and patient decisions about appropriate health care for specific clinical circumstances” as defined by the Institute of Medicine^5^. It is noted that clinical guidelines are increasingly a part of clinical practice that have potential benefits and harms; the rigorously developed evidence-based guidelines minimise possible harms, however they are merely one way of improving quality of care^6^. NICE have produced numerous documents offering guidance for the management of diabetes. Some are specifically targeted at children and young people, or pregnancy/maternity care, and thus outside the scope of this study. The relevant guidelines for adults are NG17 - Type 1 diabetes in adults: diagnosis and management^7^; and NG28 - Type 2 diabetes in adults: management^8^. In the health and social care context, quality standards set out the priority areas for quality improvement and are directed at areas where there is variation in care. Quality standards produced by NICE for the UK provide a set of statements to improve quality and information on how to measure progress^9^. They are not mandatory for implementation, but can be used to plan, commission and deliver the best possible care, to support delivering the best possible health outcomes, and are based on NICE clinical guidelines and other NICE-accredited sources^9^. The relevant quality standard is QS6 - Diabetes in adults^10^. The NICE definition of clinical audit is “a quality improvement process that seeks to improve patient care and outcomes through systematic review of care against explicit criteria and the implementation of change” ^11^. It is a technique understood globally as an effective method for evaluating health care provided by a service^12^ and a major tool for clinical governance. They can be performed as singular exercises or ongoing regular (usually at least annual) reviews and can scrutinise care nationwide (such as the National Diabetes Audit [NDA], National Adult Cardiac Surgery Audit, Paediatric Intensive Care Audit^13^ or local audits performed in trusts, hospitals or GP practices.

The NDA is a major clinical audit – defined as “a quality improvement process that seeks to improve patient care and outcomes through systematic review of care against explicit criteria and the implementation of change”^11^. NDA is one of the largest of its kind, completed every year in England and Wales since 2003. As part of its remit, it identifies frequency and levels of clinical indicators relevant to NICE guidelines. This feeds back into NICE reports on uptake and progress on its work, examining how its guidelines and targets are being implemented in the relevant clinical settings. The NDA, commissioned by the Healthcare Quality Improvement Partnership (HQIP) and delivered by NHS Digital in collaboration with Diabetes UK and Public Health England, measures the effectiveness of diabetes healthcare against NICE Clinical Guidelines and NICE Quality Standards in England and Wales. The NDA collects and analyses data for a range of stakeholders to improve the quality of services and outcomes for people with diabetes. The audit addresses these key questions: Is everyone with diabetes diagnosed and recorded on a practice diabetes register? What percentage of people registered with diabetes received the nine NICE key processes of diabetes care? What percentage of people registered with diabetes achieved NICE defined treatment targets for glucose control, blood pressure and blood cholesterol? What are the rates of acute and long-term complications (disease outcomes)?

This study investigated whether there are differences between T1DM and T2DM diabetes in the rates of uptake of relevant NICE clinical guidelines, by analysis of three years of NDA data for England and Wales. It aims to compare rates of uptake of NICE clinical processes and treatment targets between ‘Type 1’ and ‘Type 2 and other’ and consider potential explanations for any observed differences and the implications for clinical practice, diabetes management, health policy and further research. This is the first time this analysis has been performed and is being studied due to apparent differences in proportions meeting NICE treatment targets and care processes between diabetes subtypes in the NDA report^14^: understanding this is both clinically interesting and may yield important lessons for healthcare public health.

## METHODS

### National Diabetes Audit

The NDA is commissioned by the Healthcare Quality Improvement Partnership (HQIP) and delivered by NHS Digital, in collaboration with Diabetes UK and Public Health England. The NDA measures the effectiveness of diabetes healthcare against NICE Clinical Guidelines and NICE Quality Standards in England and Wales. The NDA collects and analyses data for a range of stakeholders to improve the quality of services and outcomes for people with diabetes. The audit addresses questions regarding diabetes diagnosis and recording;

- uptake of NICE recommended processes of diabetes care;
- achievement of NICE defined treatment targets (glucose control, blood pressure and blood cholesterol); and
- disease outcomes.

The NDA allows organisations to compare their outcomes with similar bodies, identify and share best practice, improve commissioning, and provide a more comprehensive picture of diabetes care and outcomes. Participating local services can benchmark their performance, identify where they are performing well, and target quality improvements. On a national level, wide participation in the audit also provides an overview of the quality of care being provided in England and Wales. NDA data is linked to Hospital Episode Statistics (HES, http://digital.nhs.uk/hes), Admitted Patient Care (APC) data, and Office for National Statistics mortality statistics data, via the NHS Digital Data Access Request Service (DARS), to provide follow up periods for complication and mortality ratio analyses. Various regular reports are produced from the NDA detailing annual information on care processes and treatment targets^3,14,15^.

### DATA

#### National Diabetes Audit data collections

The NDA team at NHS Digital kindly provided 2 bespoke cuts of data which covered a total of five NDA collection years. Where reported frequencies and proportions differ from the official NDA reports this is due to age cut-offs (this data set only includes patients aged ≥20, NDA reports cover all ages), and data for 2016-17 and 2018-19 includes rounding to the nearest 5 (i.e. 0 or 5):

- 2013-14 featuring 1 739 175 individual patients - approximately 57% of target population (identified as all patients with diabetes in England and Wales).
- 2014-15 featuring 1 871 320 individual patients - approximately 57% of target population.
- 2015-16 featuring 2 688 106 individual patients - approximately 82% of target population; this was the latest data available at the time of request.
- 2016-1 7 featuring 3 095 275 individual patients (rounded) - approximately 95% of target population
- 2017-18 featuring 3 357 055 individual patients (rounded) - approximately 98% of target population;
- Each year contained a full data set as below. Demographic and predictor variables were:
  - sex (male, female);
  - age group (in seven 10-year bands from 20-29, and then 90+ inclusive);
  - diabetes type (‘T1DM’, or ‘T2DM and Other’).

#### Outcome measures

The recommendations set out in QS6^10^ are based on public health guideline PH38^16^, and clinical guidelines NG28, NG17 and NG19^7,8,17^. Clinical care processes and treatment targets are specifically indicated by NICE and these are described in Appendix 1.

Outcome measures were six of the nine NICE diabetes clinical guideline target areas:

1. HbA1c
  - presence/absence in year of glycated haemoglobin (HbA1c) monitoring
  - whether HbA1c levels were at or below 7.5%
2. Blood pressure
  - presence/absence in year of blood pressure monitoring
  - whether blood pressure was below 140 / 80 mmHg (subsequently dropped from analysis due to error identified)
3. Cholesterol
  - the presence/absence in year of cholesterol monitoring
  - whether blood cholesterol was below 5 mmol/l
4. Urinary albumin
  - the presence/absence in year of albumin:creatinine ratio monitoring
5. Smoking review
  - the presence/absence in year of smoking status monitoring
6. Body Mass Index (BMI)
  - the presence/absence in year of Body Mass Index monitoring

The NICE care processes not included in this study were kidney function, foot exam and diabetic eye screening, as this analysis focused on outcomes addressing cardiovascular impact. The albumin:creatinine measure was included to observe whether any comparative clinical impact was observed from urine sample request.

### STATISTICAL ANALYSES

Each collection year was compared in terms of demographics and predictor: age group (in 10-year bands from 20-29 to 90+ inclusive), sex, and diabetes type (T1DM or T2DM and other). Each NICE care process and treatment target included was described for ‘Type 1’ and ‘Type 2 and Other’ for each audit year. Where patients did not meet the monitoring criteria for HbA1c, BP or cholesterol, they were excluded from the denominator for the relevant treatment target level statistics.

Univariable logistic regression was performed to explore crude associations between diabetes type and each outcome variable. Multivariable logistic regression was performed to examine associations between the outcome variables and diabetes type adjusting for age group and sex. Each logistic regression was performed on a dataset formed from the relevant collection year, using as reference categories T1DM, male sex, age group 20-29. These were chosen as reference categories as they are often reported to be the poorest engagers with healthcare^18-21^. In the case of those variables where the outcome variable was meeting a target level of a biometric (HbA1c, BP, cholesterol), observations were excluded in the year where the monitoring was signalled not to have taken place.

The analysis using univariable and multivariable regression models allowed for comparisons to highlight potential effects of age and/or sex on the observed associations.

Statistical analysis was performed using STATA version 14.2 software.

### PATIENT AND PUBLIC INVOLVEMENT

We did not directly include PPI in this study, but the database used in the study was developed with PPI and is updated by a committee that includes patient representatives.

## RESULTS

### Patients in National Diabetes Audit

Table 1 below shows how numbers have increased annually in the latest three year’s data for NDA, and the relative amounts of ‘Type 1’ and ‘Type 2 and other’ have slowly moved toward an increased proportion of ‘Type 2 and other’. T2DM patients outnumber T1DM in the audit by about 10 to 1, which is consistent with current understanding of UK and global prevalence rates^22^.

**Table 1.**
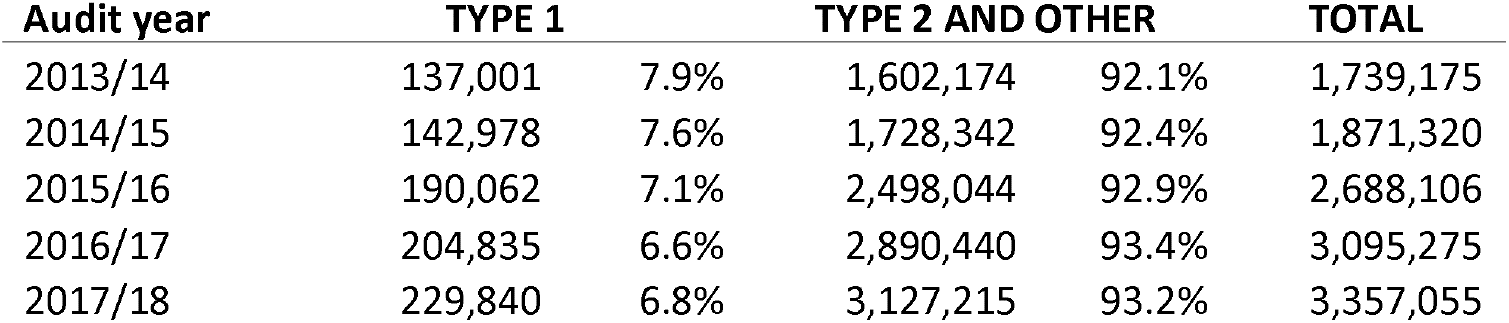
Number of patients and relative proportions of each diabetes type reported in the National Diabetes Audit of the past five annual collections

Table 2 below shows the male/female frequencies and proportions for each collection year by diabetes type. It shows a very high level of consistency between the three collection years analysed, despite different levels of coverage of the diabetic population and submission rates to the audit. There is also consistency in terms of the proportions of T1DM and T2DM engaging with the audit, again despite the different submission rates and penetration. The increase in actual numbers displays how submissions to the audit have increased over time.

**Table 2.** Numbers and proportions of males and females by diabetes type in each National Diabetes Audit collection year 2013-14, 2014-15, 2015-16, 2016-17, 2017-18

| Audit year | Male |  | Female |  | TOTAL |
| --- | --- | --- | --- | --- | --- |
|  | TYPE 1 |  |  |  |  |
| 2013/14 | 77,782 | 56.8% | 59,219 | 43.2% | 137,001 |
| 2014/15 | 81,188 | 56.8% | 61,790 | 43.2% | 142,978 |
| 2015/16 | 108,305 | 57.0% | 81,757 | 43.0% | 190,062 |
| 2016/17 | 116,930 | 57.0% | 87,905 | 43.0% | 204,835 |
| 2017/18 | 131,240 | 57.1% | 98,600 | 42.9% | 229,840 |
|  | TYPE 2 AND OTHER |  |  |  |  |
| 2013/14 | 893,120 | 55.7% | 709,054 | 44.3% | 1,602,174 |
| 2014/15 | 962,139 | 55.7% | 766,203 | 44.3% | 1,728,342 |
| 2015/16 | 1,389,643 | 55.6% | 1,108,401 | 44.4% | 2,498,044 |
| 2016/17 | 1,611,875 | 55.8% | 1,278,565 | 44.2% | 2,890,440 |
| 2017/18 | 1,743,220 | 55.7% | 1,383,995 | 44.3% | 3,127,215 |

Figure 1 below profiles the age group distributions of the three NDA collection years according to diabetes type. There is a very high level of consistency across the five collection years according to each diabetes type. The marked difference in age distributions between T1DM and T2DM is predictable given the natural history of each condition, age of onset, diagnosis, and premature mortality profile. The large increases in number of patients included in 2015-16 and 2016-17 did not change the appearance of either diabetes type age profile, increasing face validity of the prior datasets.

**Figure 1.**
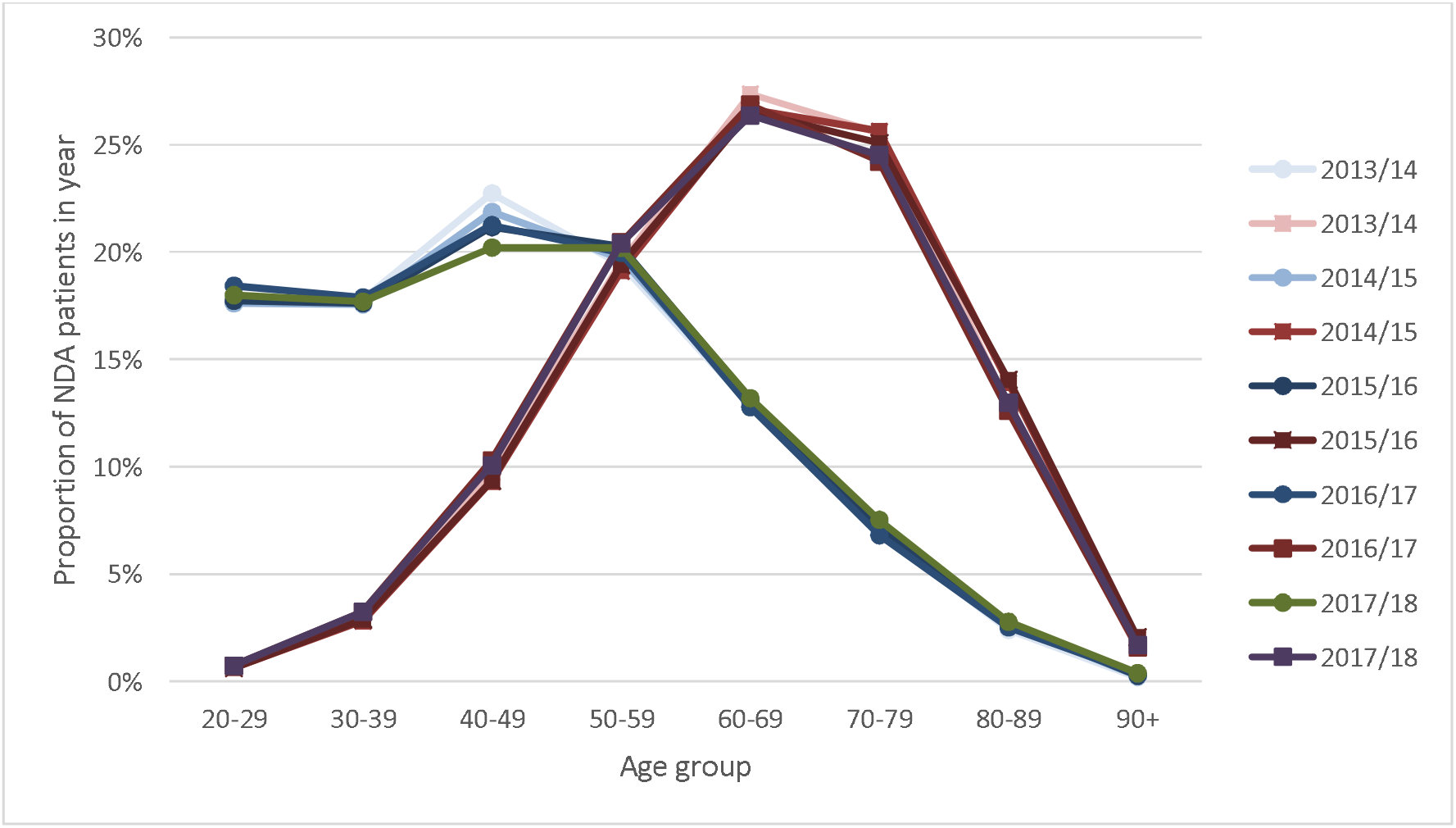
Chart showing the age distributions (from 20 years and up) of the T1DM (round markers) and T2DM (square markers) populations profiled by the National Diabetes Audits of 2013-14, 2014-15, 2015-16, 2016-17 and 2017-18

### Trends in progress against NICE targets for Type 1 and Type 2 diabetes

Data presented in Table 3 shows how the audit years have been relatively consistent in terms of proportions of Type 1 and Type 2 patients meeting NICE clinical processes and treatment targets. The most met care process for Type 1 diabetes was blood pressure monitoring (87% in 2013-14, increasing monotonically to 91% in 2017-18). The least commonly met treatment target for Type 1 was HbA1c<=7.5% (29.1% in 2013-14, to 30.1% in 2017-18, with highest value of 30.8% in 2016-17). The most met care process for patients with Type 2 diabetes was blood pressure monitoring (92.9% in 2013-14, to a high of 95.3% in 2016-17 and 2017-18). The least commonly met treatment target for Type 2 and other was HbA1c<=7.5% (65.4% in 2013-14 and in 2017-18, with highest value of 66.8% in 2016-17).

**Table 3.**
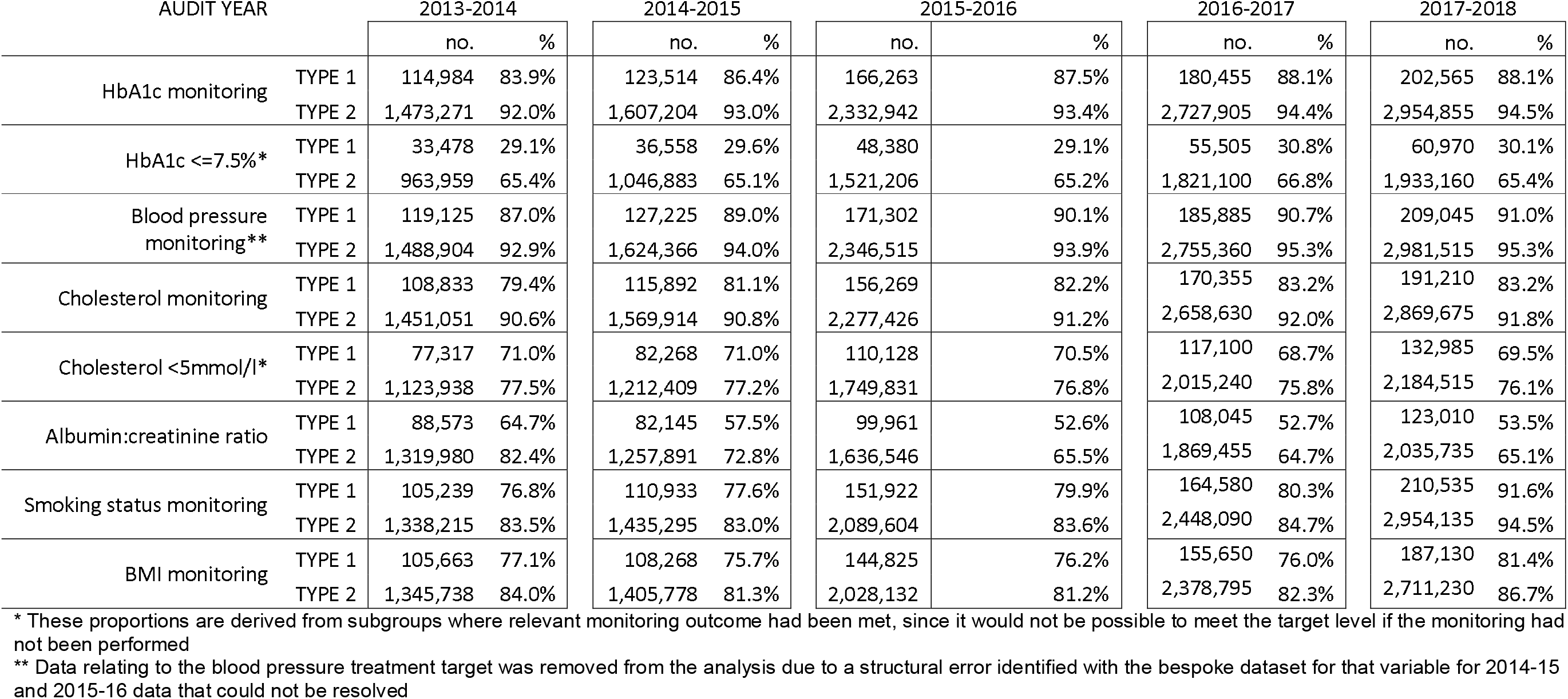
Numbers and proportions receiving NICE processes and meeting NICE targets by diabetes

| AUDIT YEAR |  | 2013-2014 |  | 2014-2015 |  | 2015-2016 |  | 2016-2017 |  | 2017-2018 |  |
| --- | --- | --- | --- | --- | --- | --- | --- | --- | --- | --- | --- |
|  |  | no. | % | no. | % | no. | % | no. | % | no. | % |
| HbA1c monitoring | TYPE 1 | 114,984 | 83.9% | 123,514 | 86.4% | 166,263 | 87.5% | 180,455 | 88.1% | 202,565 | 88.1% |
|  | TYPE 2 | 1,473,271 | 92.0% | 1,607,204 | 93.0% | 2,332,942 | 93.4% | 2,727,905 | 94.4% | 2,954,855 | 94.5% |
| HbA1c <=7.5%* | TYPE 1 | 33,478 | 29.1% | 36,558 | 29.6% | 48,380 | 29.1% | 55,505 | 30.8% | 60,970 | 30.1% |
|  | TYPE 2 | 963,959 | 65.4% | 1,046,883 | 65.1% | 1,521,206 | 65.2% | 1,821,100 | 66.8% | 1,933,160 | 65.4% |
| Blood pressure monitoring** | TYPE 1 | 119,125 | 87.0% | 127,225 | 89.0% | 171,302 | 90.1% | 185,885 | 90.7% | 209,045 | 91.0% |
|  | TYPE 2 | 1,488,904 | 92.9% | 1,624,366 | 94.0% | 2,346,515 | 93.9% | 2,755,360 | 95.3% | 2,981,515 | 95.3% |
| Cholesterol monitoring | TYPE 1 | 108,833 | 79.4% | 115,892 | 81.1% | 156,269 | 82.2% | 170,355 | 83.2% | 191,210 | 83.2% |
|  | TYPE 2 | 1,451,051 | 90.6% | 1,569,914 | 90.8% | 2,277,426 | 91.2% | 2,658,630 | 92.0% | 2,869,675 | 91.8% |
| Cholesterol <5mmol/l* | TYPE 1 | 77,317 | 71.0% | 82,268 | 71.0% | 110,128 | 70.5% | 117,100 | 68.7% | 132,985 | 69.5% |
|  | TYPE 2 | 1,123,938 | 77.5% | 1,212,409 | 77.2% | 1,749,831 | 76.8% | 2,015,240 | 75.8% | 2,184,515 | 76.1% |
| Albumin:creatinine ratio | TYPE 1 | 88,573 | 64.7% | 82,145 | 57.5% | 99,961 | 52.6% | 108,045 | 52.7% | 123,010 | 53.5% |
|  | TYPE 2 | 1,319,980 | 82.4% | 1,257,891 | 72.8% | 1,636,546 | 65.5% | 1,869,455 | 64.7% | 2,035,735 | 65.1% |
| Smoking status monitoring | TYPE 1 | 105,239 | 76.8% | 110,933 | 77.6% | 151,922 | 79.9% | 164,580 | 80.3% | 210,535 | 91.6% |
|  | TYPE 2 | 1,338,215 | 83.5% | 1,435,295 | 83.0% | 2,089,604 | 83.6% | 2,448,090 | 84.7% | 2,954,135 | 94.5% |
| BMI monitoring | TYPE 1 | 105,663 | 77.1% | 108,268 | 75.7% | 144,825 | 76.2% | 155,650 | 76.0% | 187,130 | 81.4% |
|  | TYPE 2 | 1,345,738 | 84.0% | 1,405,778 | 81.3% | 2,028,132 | 81.2% | 2,378,795 | 82.3% | 2,711,230 | 86.7% |

\* These proportions are derived from subgroups where relevant monitoring outcome had been met, since it would not be possible to meet the target level if the monitoring had not been performed
\*\* Data relating to the blood pressure treatment target was removed from the analysis due to a structural error identified with the bespoke dataset for that variable for 2014-15 and 2015-16 data that could not be resolved

The chart below (figure 2) shows that across the three profiled audit years, there have been general trends in small increases in progress toward meeting NICE clinical processes. A clear exception is visible in the monitoring of albumin:creatinine ratio. This has seen a marked decrease in both Type 1 and Type 2 patient groups. The treatment targets around glycaemic control, blood pressure and cholesterol have remained stable over the three audit years. In both patient groups, the most delivered NICE treatment targets were monitoring of HbA1c and blood pressure. In both T1DM and T2DM the most frequently missed treatment target was HbA1c<=7.5%. However, in Type 1 there were two outcomes mostly <60% (HbA1c<=7.5%, and albumin:creatinine ratio). Treatment target of cholesterol <5mmol/l was lower for Type 1s than Type 2s however there were lower rates of cholesterol monitoring overall.

**Figure 2.**
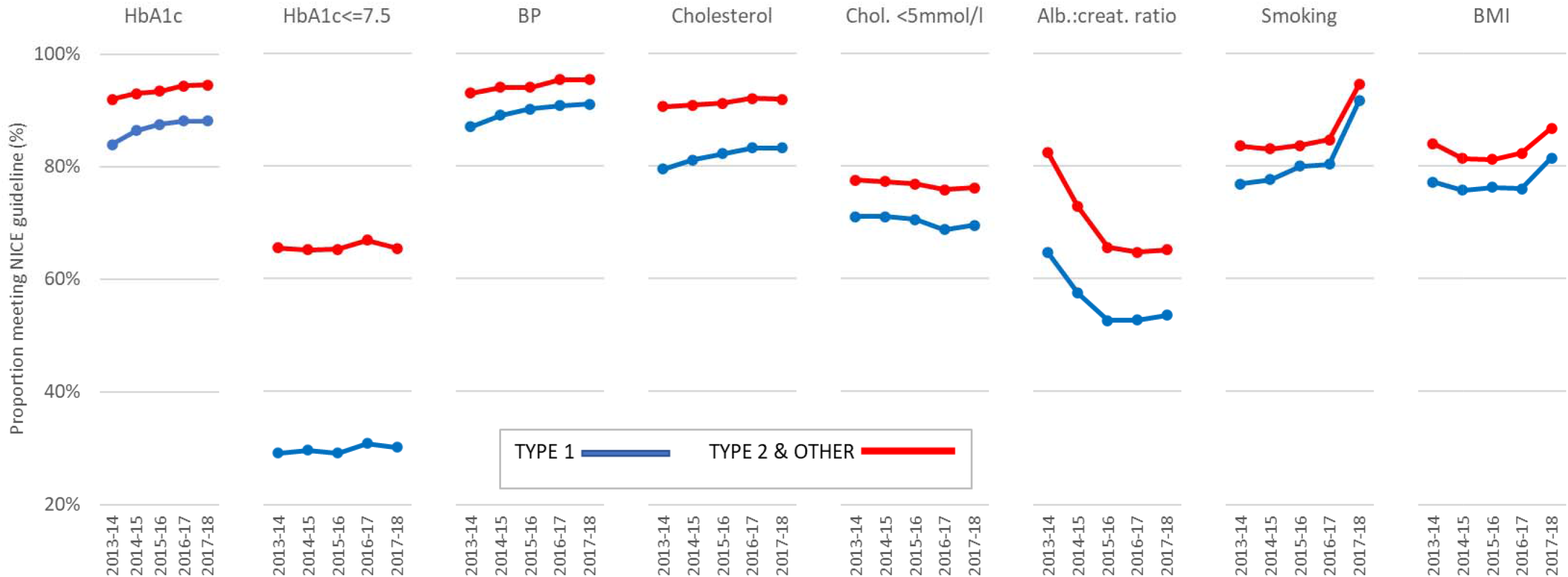
Chart showing trend from 2013-14 through to 2015-16 in proportion of Type 1 and Type 2 diabetes patients included in National Diabetes Audit meeting the relevant NICE treatment target or care process

### Associations between patient characteristics and NICE processes and treatment targets

Logistic regression analysis was performed on the outcome variables pertaining to the NICE care processes and treatment targets utilising diabetes type as the predictor variable before (crude rate) and after (adjusted rate) controlling for age and sex: a profile of odds ratios is shown in Table 4 below. T1DM was used as the reference category. Thus, the ORs presented shows the odds of a patient with T2DM meeting the relevant treatment target versus a patient with T1DM: for example, in 2015-16 the odds of a T2DM patient meeting their HbA1c<=7.5% target was 3.43 (95%CI 3.39-3.47) times that of a T1DM patient (adjusted for age and sex). Reference categories for age group and sex were ‘20-29 years’ and ‘male’ respectively.

**Table 4.** Odds Ratios (95% Confidence Intervals) of Type 2 Diabetes Mellitus patients meeting NICE clinical processes and treatment targets compared with Type 1 (reference category) in five recent National Diabetes Audit collections^ (crude, and adjusted for age group and sex)

|  |  | 2013-14 | 2014-15 | 2015-16 | 2016-17 | 2017-18 |
| --- | --- | --- | --- | --- | --- | --- |
| Glycated haemoglobin (HbA1c) monitoring | univariable | 2.19 (2.15-2.22) | 2.09 (2.06-2.12) | 2.02 (1.99-2.05) | 2.27 (2.24-2.30) | 2.31 (2.28-2.34) |
|  | multivariable* | 1.25 (1.23-1.28) | 1.17 (1.14-1.19) | 1.09 (1.08-1.11) | 1.29 (1.26-1.31) | 1.27 (1.25-1.29) |
| HbA1c ≤7.5% | univariable | 4.61 (4.55-4.69) | 4.44 (4.39-4.50) | 4.57 (4.52-4.62) | 4.58 (4.54-4.63) | 4.48 (4.44-4.53) |
|  | multivariable* | 3.42 (3.37-3.46) | 3.31 (3.26-3.35) | 3.43 (3.39-3.47) | 3.19 (3.15-3.22) | 3.21 (3.18-3.24) |
| Blood pressure monitoring** | univariable | 1.97 (1.94-2.00) | 1.93 (1.90-1.97) | 1.70 (1.67-1.72) | 2.08 (2.05-2.11) | 2.04 (2.00-2.07) |
|  | multivariable* | 1.13 (1.11-1.15) | 1.05 (1.03-1.07) | 0.89 (0.87-0.90) | 1.12 (1.10-1.14) | 1.03 (1.02-1.05) |
| Cholesterol monitoring | univariable | 2.49 (2.45-2.52) | 2.32 (2.28-2.35) | 2.23 (2.20-2.26) | 2.32 (2.29-2.35) | 2.25 (2.23-2.28) |
|  | multivariable* | 1.38 (1.36-1.40) | 1.32 (1.30-1.35) | 1.26 (1.24-1.28) | 1.38 (1.36-1.40) | 1.33 (1.31-1.34) |
| Cholesterol <5mmol/l | univariable | 1.40 (1.38-1.42) | 1.39 (1.37-1.40) | 1.39 (1.37-1.41) | 1.73 (1.71-1.74) | 1.69 (1.67-1.70) |
|  | multivariable* | 0.97 (0.96-0.99) | 0.97 (0.95-0.98) | 0.97 (0.96-0.98) | 1.12 (1.10-1.13) | 1.09 (1.08-1.10) |
| Smoking status monitoring | univariable | 1.53 (1.51-1.55) | 1.41 (1.40-1.43) | 1.28 (1.27-1.30) | 1.35 (1.34-1.37) | 1.57 (1.54-1.59) |
|  | multivariable* | 1.32 (1.30-1.34) | 1.27 (1.25-1.29) | 1.14 (1.13-1.16) | 1.27 (1.25-1.29) | 1.16 (1.14-1.18) |
| Albumin:creatinine ratio monitoring | univariable | 2.56 (2.53-2.59) | 1.98 (1.96-2.00) | 1.71 (1.70-1.73) | 1.61 (1.63-1.66) | 1.62 (1.61-1.63) |
|  | multivariable* | 1.43 (1.41-1.45) | 1.18 (1.17-1.20) | 1.09 (1.08-1.10) | 1.09 (1.08-1.10) | 1.10 (1.09-1.11) |
| Body Mass Index monitoring | univariable | 1.56 (1.54-1.58) | 1.40 (1.38-1.42) | 1.35 (1.33-1.36) | 1.47 (1.45-1.48) | 1.49 (1.47-1.50) |
|  | multivariable* | 1.22 (1.20-1.23) | 1.17 (1.15-1.19) | 1.11 (1.10-1.13) | 1.26 (1.25-1.28) | 1.20 (1.19-1.22) |

\* adjusted for age group and sex
\*\* Data relating to the blood pressure treatment target was removed from the analysis due to a structural error identified with the bespoke dataset for that variable for 2014-15 and 2015-16 data that could not be resolved
All OR presented are significant p<.0001

Both crude and adjusted ORs reported for T2DM are mostly greater than 1 (see Table 4), suggesting that T2DM patients achieve the NICE guidelines more reliably than T1DM. As noted, this is most marked in the HbA1c<=7.5% outcome, where the odds of T2DM patients meeting their glycated haemoglobin targets were more than 3 times the odds of T1DM patients (ORs between 3.19 (95% CIs 3.39-3.47) in 2016-17 and 3.43 (95% CIs 3.39-3.47) in 2015-16) after adjusting for age and sex.

There is consistency across the five audit years; however, there is a general trend in the observed ORs across the five years for many of the outcome variables to approach the baseline, suggesting the differences between T1DM and T2DM groups meeting NICE processes and treatment targets were gradually narrowing. The only ORs to increase slightly in the final year were the HbA1c<=7.5% and albumin:creatinine ratio outcomes. The increases in sample size (particularly) from 2014-15 to 2015-16 (1.9 million to 2.7 million patients submitted to the audit) were the most notable temporal changes.

ORs were <1 for treatment targets for cholesterol <5mmol/l, suggesting once age and sex are considered patients with T1DM were more likely to meet these targets than T2DM. In each audit year T2DM is more likely to register cholesterol <5mmol/l until confounders are addressed. This indicates that T2DM patients are more likely than T1DM to report lower cholesterol until age is adjusted for, whereby the difference disappears, and Type 1 reports slightly greater achievement of low cholesterol. OR is also <1 in 2015-16 for blood pressure monitoring, showing that after adjusting for age and sex, T1DM have moved from being less likely to be BP monitored in 2013-14 and 2014-15 to more likely to be monitored than T2DM in 2015-16. However, this OR returns to >1 in 2016-1 7 and 2017-18.

## DISCUSSION

Clear differences are observed between ‘Type 1’ and ‘Type 2 and other’ diabetes groups regarding NICE care processes and treatment targets within each audit year (p<.001). The critical glycaemic control guideline (HbA1c<=7.5%) is over 3 times the odds of being met by T2DM compared to T1DM, consistent over the five years reported: characterising a key clinical difference between the two DM types. In general, T2DM patients have been more successful in meeting these care processes and targets. Significance levels were consistently very high resulting from large samples: more emphasis is given to ORs and 95% confidence intervals when considering the associations. In 2017-18, top proportions meeting NICE recommendations for T1DM and T2DM were 91.4% - for providing smoking status - and 95.3% for BP monitoring respectively. Proportions of patients receiving NICE recommended clinical processes (e.g. monitoring of HbA1c and blood pressure) increased from 2013 to 2016, but no observable progress was made for biometric treatment targets (e.g. HbA1c<=7.5%). Adjusting for age when looking at differences between T1DM and T2DM is clearly important. Performing multivariable logistic regressions made no associations less significant, but in some cases reversed the direction of the association. This was observed with serum cholesterol <5mmol/l in each audit year: the crude rate suggested T2DM meets the target more frequently, but the adjusted rate showed that T1DM meets lower cholesterol targets more readily after controlling for age and sex effects.

The increase in proportions of patients receiving NICE recommended clinical processes but not biometric treatment targets may suggest that clinical guidelines are becoming more successful at driving clinician process behaviour, but this is not translating into improved patients’ self-management of their diabetes, irrespective of diabetes type.

Blood pressure monitoring is apparently well embedded in clinical practice and well tolerated by most patients as >90% of both T1DM and T2DM are receiving. It also shows that high levels of engagement occur for those patients registered on the audit, since these proportions of patients are contacting relevant services within 12 months.

After controlling for age adjusted rate showed that T1DM meets lower cholesterol targets more readily than T2DM, which may reflect lifestyle factors associated with T2DM such as diet and obesity^23^.

The HbA1c<=7.5% success of T2DM compared to T1DM underpins the increased mortality risk with T1DM, with the higher rates of hyperglycaemia associated with CVD and premature death^24^. Challenges around monitoring of blood glucose and the administration of insulin for this group

### Strengths and limitations of this study

There are legitimate concerns about misclassification in this study on two points: the clinical diagnoses of diabetes type^25,26^ and the actual category (met/not met) in the clinical audit response. Recent literature has questioned the way clinicians historically diagnosed/classified diabetes^27-29^ suggesting differentiation between T1DM and T2DM may be less clear than previously thought. Studies investigating the accuracy of diabetes diagnosis in UK primary care^30,31^ have identified incorrect diagnoses, misclassification of diabetes type, and use of ambiguous Read codes potentially introducing errors into the dataset. T2DM patients may be more likely to be “healthcare engagers” and may therefore be a self-selecting group. T1DM patients cannot survive without insulin and will therefore be highly likely to be registered with primary or secondary care. Conversely, T1DM patients may feature a higher proportion who resist engaging with health professions generally. Potential sampling bias also relates to those GP practices not submitting. Concerns there may have been bias from services covering patients not submitting data to the NDA (40% in 2013/14 and 2014/15, and 16% in 2015/16) compromising validity of data were allayed by consistency observed in the 2016-17 and 2017-18 data, where proportion of non-submission was 5% and 3% respectively. NDA reports include CCG mapping which allows some quality assurance, but there may have been issues with non-submitting practices and clinics nationally. Not all NICE recommendation data available through the NDA was utilised and it is possible that these variables may have shown a different pattern of outcomes, potentially varying some of the general conclusions. Additional confounders may have influenced the findings, such as ethnicity, deprivation decile and access to services. Their absence from the dataset and analysis is acknowledged.

Statistics were robust and a large sample size means that confidence intervals and p values will be precise. Consistency of measured patient group characteristics supports the reliability of the audit process. Data completeness flaws characterises a typical challenge associated with using routine data for epidemiological research. Over a million people in the UK, mostly with Type 2 diabetes, are undiagnosed and unregistered^2^. It is not known how effectively this unregistered group would attain NICE treatment targets and care processes if they engaged with services. The annual repeat of the NDA has yielded more than a decade of historical data and continuously improved data architecture and collection methodology, allowing analysis of trends and an increasingly useful dataset. Validity between audit years and analysis that supports comparability can be explored further in the NDA Data Quality Statements^32,33^. The final manuscript was assessed using the STROBE guidelines for reporting observational studies^34^ and compliance with this checklist can be seen in web appendix.

An error was identified in the data file from NHS Digital meaning data pertaining to blood pressure targets were excluded from the results presented in this paper.

### Comparison with other studies

Comparisons of strengths and limitations with other studies is challenging since there is little published literature based on NDA data. Following up previous research, future studies might profile T1DM patients who successfully meet the NICE targets, particularly regarding demographics, locations, and other markers. Identifying any relationship or commonalities with target compliant T2DM patients could recognise which groups respond best, which groups may require additional or different intervention to meet NICE guidelines. It would also be vital to look at clinical factors that may improve chance of successfully meeting NICE targets: e.g. clinician communication style, method, timing of interventions and particularly setting (primary or secondary care).

Blood pressure levels and cholesterol levels targets were more readily met by T1DM patients as shown by ORs <1 after controlling for age, possibly highlighting clinical precursors of T2DM with concurrent obesity and insufficient physical activity; or reflecting impact of medication if levels of statins prescribing differs between T1DM and T2DM. Literature exploring this issue is absent, and thus a potential focus for further research.

Future studies could expand the current methodology to explore all 9 of the NICE diabetes targets individually, collectively, and clustered according to disease focus (i.e. CVD, CKD, sensory organs, foot problems) or data collection modality (blood etc.), to allow a comprehensive understanding of factors improving engagement and compliance and provide a better picture of NICE guideline impact for this increasingly urgent and important disease.

### Conclusions and public health implications

The NDA is a powerful and effective way of monitoring epidemiology of diabetes in England and Wales, and for monitoring the uptake and impact of relevant clinical guidelines. NDA submission became mandatory for primary care and specialist units in 2016-17, underpinning the current 98% coverage.

Diabetes type is strongly associated with rates of uptake of NICE clinical guidelines after controlling for age and sex. The extremely high number of patients included in the NDA provides robust confidence in the strength of these associations. T1DM patients do not appear to meet NICE treatment targets as readily as T2DM, and where they do, this may be more related to problems for T2DM patients associated with their wider health and lifestyle factors such as issues with diet, exercise, and obesity/overweight.

HbA1c (as a measure of medium-term glycaemic control) is key for CVD and other diabetic complications, and T1DM patients especially struggle to meet the targets. Adequate consideration should be applied to this situation: including options for setting different HbA1c targets for T1DM and T2DM, especially as persistent failure to achieve the target level may be demotivating for the patient and thus clinically counter-productive. Cost-effective measures providing additional support or technology for T1DM patients in managing their HbA1c specifically, or managing blood glucose levels acutely, such as continuous glucose monitoring could be another solution. The recommendations published in NG17 (NICE 2015a) require close monitoring of their impact on the NICE treatment targets and clinical processes profiled in the NDA. These mainly focus on structured education programmes, supporting adults for improved HbA1c management, specific blood glucose targets at certain times of day, etc. (see Appendix 1). Future NDA analyses could also examine the impact of expansion of continuous glucose monitoring and insulin pumps and help make assessments to determine future clinical systems and policy decision-making. As part of this future consideration, questions must be addressed regarding what appropriate care is, especially for patients requiring complex and expensive therapeutic interventions at advanced ages.

## Data Availability

Raw data were generated at NHS Digital. Derived data supporting the findings of this study are available from the corresponding author RCH on request.

## CONTRIBUTORS

Dr Anna Richards, Devon County Council

Dr Catherine Falconer, Glos Hospitals NHS Trust

Peter Knighton, NHS Digital

Prof Chris J Hyde, University of Exeter

## ETHICAL APPROVAL

The use of aggregated data from NDA did not necessitate ethics committee approval.

## DATA SHARING

NHS Digital provided two bespoke datasets based on aggregated data from the NDA collection years: Packet 1 2013-14, 2014-15, 2015-16 & Packet 2 2016-17 and 2017-18.

## TRANSPARENCY

The lead author* affirms that this manuscript is an honest, accurate, and transparent account of the study being reported; that no important aspects of the study have been omitted; and that any discrepancies from the study as planned (and, if relevant, registered) have been explained.

